# Accurate classification of secondary progression in multiple sclerosis

**DOI:** 10.1101/2020.07.09.20149674

**Authors:** Ryan Ramanujam, Feng Zhu, Katharina Fink, Virginija Danylaitė Karrenbauer, Johannes Lorscheider, Pascal Benkert, Elaine Kingwell, Helen Tremlett, Jan Hillert, Ali Manouchehrinia, The BeAMS Study group

## Abstract

Transition from a relapsing-remitting to the secondary progressive phenotype is an important milestone in the clinical evolution of multiple sclerosis. In the absence of reliable imaging or biological markers of phenotype transition, assignment of current phenotype status relies on retrospective evaluation of the medical history of an individual. Here, we sought to determine if demographic and clinical information from multiple sclerosis patients can be used to accurately assign current disease phenotypes: either relapsing-remitting or secondary progressive status. Data from the most recent clinical visit of 14,387 multiple sclerosis patients were extracted from the Swedish Multiple Sclerosis Registry. Decision trees based on sex, symptom onset age, Expanded Disability Scale Status score, and age & disease duration at the most recent clinic visit, were examined to build a classifier to determine disease phenotype. Validation was conducted using an independent cohort of multiple sclerosis patients from British Columbia, Canada, and a previously published classifier to assign phenotype was also tested. Clinical records of 100 randomly selected patients were used to manually categorize phenotype by three independent neurologists. A decision tree (the classifier) containing only most recently available disability score and age obtained 89.3% (95% confidence intervals (CI): 88.8% to 89.8%) classification accuracy, defined as concordance with the latest reported status in the registry. Replication in an independent cohort from British Columbia resulted in 82.0% (95%CI: 81.0% to 83.1%) accuracy. A previously published classification algorithm with slight modifications achieved 77.8% (95%CI: 77.1% to 78.4%) accuracy when assigning disease phenotype. With complete patient history data, three neurologists obtained 84.7% accuracy on average compared with 85 for the classifier using the same data. The model is easily interpretable and could allow research studies and randomized clinical trials to estimate the probability of patients having already reached the secondary progressive stage when they have not yet been retrospectively assigned this status, and to standardize definitions of disease phenotype across different cohorts. Clinically, this model could assist neurologists by providing additional information about the probability of having secondary progressive disease. This could also benefit patients who may be introduced to new therapies targeting progressive multiple sclerosis.

## Introduction

Multiple sclerosis is a chronic demyelinating disorder, most often of a relapsing-remitting (RR) course. After many years, the disease course typically converts to a secondary progressive (SP) phase wherein accumulation of irreversible disability occurs and the disease progresses steadily throughout a patient’s remaining life, often in the absence of clinical relapses. (Lublin *et al*., 2014) The average time from a RR disease onset to transition to SP disease is approximately 20 years. (Tremlett *et al*., 2010) There are important clinical implications when a patient has reached SP multiple sclerosis, since most disease modifying drugs (DMDs) are indicated during the RR phase of multiple sclerosis. (Scalfari *et al*., 2014) DMDs’ efficacies also appear to wane as a person ages and the SP phase is reached. (Weideman *et al*., 2017)

The most common method of assessing the time at which the patient has transitioned to the SP phase is a retrospective clinical review of a patient’s medical history, including the expanded disability status scale (EDSS) scores (Kurtzke, 1983) over time. However, this approach may vary among clinicians or countries with different assessment criteria. Furthermore, neurologists may feel hesitant to make such an irreversible determination early, or at the time of transition, as an assignment of a SP course may render patients with limited DMD options. An objective measure of transition to SP multiple sclerosis that relies on basic clinical measurements would potentially benefit both clinicians and researchers. (T. *et al*., 2013) In the clinic, such a tool could provide a complimentary metric to assist in decision-making and reinforce clinical assessment. This tool could also benefit clinical research by creating a uniform basis for unbiased classification, thereby minimizing variation between and within studies.

We used a large pool of patients with known disease phenotype and basic clinical variables in order to build and verify a classifier. We included validation from an independent cohort, and comparisons to existing methods of assigning SP disease status.

## Materials and methods

### Patient materials

Multiple sclerosis patients with a relapsing remitting (RR) disease course at multiple sclerosis symptom onset (RR-onset) and available information on date of birth, date multiple sclerosis symptom onset, sex, year of SP transition (if applicable) and the date and score of the most recent EDSS (n=14,387) were extracted from the Swedish multiple sclerosis Registry (SMSreg, hereafter referred to as the “Swedish cohort”) (Andersen, 2012). For the Swedish cohort, the SP transition date is assigned retrospectively by the attending neurologist during a clinical visit based on international consensus. (Lublin and Reingold, 1996) .The cohort was used to build the classifier.

A cohort of 5,431 RR-onset multiple sclerosis patients from British Columbia, Canada (hereafter referred to as the “Canadian cohort”) was used to validate the classifier. This cohort has been previously described (Tremlett *et al*., 2008; Zhang *et al*., 2015) and was selected because similar information was available, including the assignment of the SP transition date.

### Construction of decision tree classifier

Several types of classification techniques including support vector machines, random forest and logistic regression model were considered. Decision trees were ultimately selected as they generate very clear rules which are easy to interpret, and can be readily applied in clinical practice. (Breiman *et al*., 1984) When assessing a patient’s clinical course, transparency to the underlying model decisions is preferred since the relevant factors can be easily confirmed manually. To benchmark the decision tree results, an alternative model was created by logistic regression using the same data as the final decision tree. Logistic regression was chosen due to ease of use, interpretability of results, and scaling via the logit function from 0 to 1.

The recursive partitioning (rPART) method (Breiman *et al*., 1984) was used to identify the optimal split of the data that would best classify the patients into the two phenotypes - RR and SP. In the first instance, four fully grown decision tree classifiers were developed using combinations of age at the most recently available EDSS assessment, EDSS score, sex, age at multiple sclerosis symptom onset and disease duration (from symptom onset) at the EDSS assessment. Variables that did not affect the classification accuracy were then removed to simplify the models. The decision tree classifier was then pruned to its simplest state by setting the complexity parameter to that of the tree with the smallest cross-validation error. The complexity parameter is the minimum improvement in the model needed in each node. We then calculated the accuracy, sensitivity, specificity, positive predictive value (PPV), and negative predictive value (NPV) of the decision tree classifier for predicting disease phenotype (RR vs. SP) at the time of the most recently available EDSS score. The decision tree classifier is a cross-sectional

### Comparison with MSBase SP algorithm

A comparison with an existing method of estimating disease status was conducted (Lorscheider *et al*., 2016). Derived from data extracted from the MSBase Registry, a large international observational multiple sclerosis collaboration, this algorithm is based on longitudinal data for each patient. The MSBase algorithm assigns conversion to SP multiple sclerosis if the following criteria are met: At least a one point increase on the EDSS for patients with an EDSS <6, and at least a 0.5 points increase for patients with an EDSS >=6, in the absence of a clinical relapse. In addition, an EDSS >=4 must be reached, and a pyramidal Functional System (FS) score of 2 or above, both confirmed at a second visit at least 3 months later (confirmed EDSS progression). In the original work, (Lorscheider *et al*., 2016) this definition achieved 87% diagnostic accuracy (compared with a consensus diagnosis of 3 multiple sclerosis neurologists) and was able to detect SP multiple sclerosis more than 3 years earlier than the physicians’ clinical assessment (using information from the same database). In the present study, this algorithm was adapted to ignore the FS Scores criterion (due to lack of availability in our data). We expect, in practice, that this adaptation should have a minimal effect on score accuracy. Further, many MS clinical databases worldwide do not routinely collect the FS sub-scores.

### Comparison with clinical evaluations

Three multiple sclerosis neurologists from the Karolinska University Hospital, Sweden (KF, JH and VD), independently and blindly reviewed the clinical records from 100 randomly chosen patients with RR onset to determine how clinical assessments compared with decision tree classifier. In the first instance, two of these neurologists classified patients using only the variables at the latest visit which were included in the decision tree classifier. Then all three neurologists repeated the classifications by using complete patient clinical records including all recorded patient visits with EDSS scores, relapses, etc.

### Comparison of time to SP conversion between different methods of classification

To compare average rates of conversion to SP multiple sclerosis between the different methods of estimating disease status, Kaplan-Meier plots were utilized with the time to SP assessed from birth as well as from multiple sclerosis symptom onset. The tree classifier outputs constructed here and predictions from the MSBase SP algorithm [12] were used and compared to the phenotype labels assigned by neurologists in the registry.

The software that was used to analyze data included R version 3.2.3 (Team, 2014) and the packages “e1071”, “party”, “rpart”, “rpart.plot” and “partykit”. Ethical permission for the study was granted by the Stockholm Regional Ethical Committee and the University of British Columbia’s Clinical Research Ethics Board.

### Data availability

The Swedish data related to the current article are available from Jan Hillert, Karolinska Institutet. To be able to share data from the Swedish multiple sclerosis registry, a data transfer agreement along with appropriate ethical permissions need to be obtained between Karolinska Institutet and the institution requesting data access. This is in accordance with the data protection legislation in Europe (General Data Protection Regulation [GDPR]). Persons interested in obtaining access to the data should contact Ali Manouchehrinia.

## Results

### Study population

In total, 14,387 patients were included in the Swedish cohort of which 71.8% were female; the average age at onset of multiple sclerosis was 32.4 years (Standard deviation (SD) ±10.2). Mean age at the most recent MS clinic visit with an EDSS score was 48.6 years (SD ±12.9) and median disease duration was 14.0 years (Interquartile range: 7.0 to 23.0). By the date of data extraction (February 2019), 68% of the patients remained in the RR phase and 32% had transitioned to SPMS (Table 1).

**Table 1:**
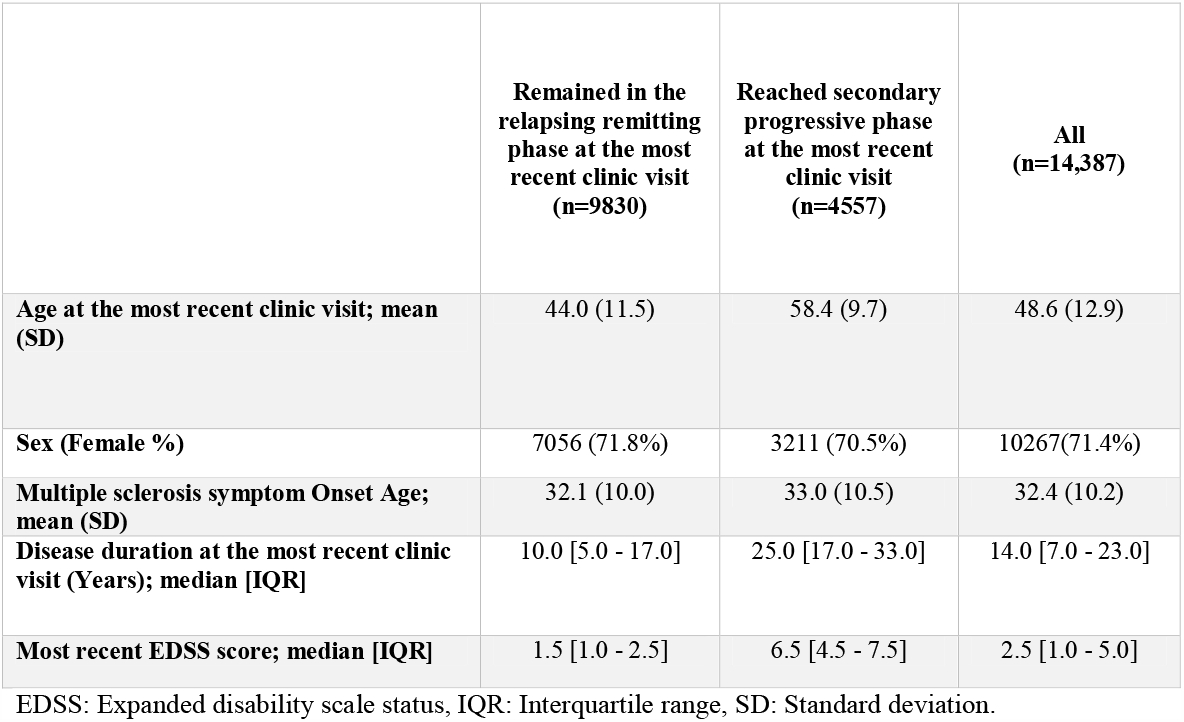
Characteristics of the Swedish cohort used to build the decision tree classifier.

|  | **Remained in the relapsing remitting phase at the most recent clinic visit (n=9830)** | **Reached secondary progressive phase at the most recent clinic visit (n=4557)** | **All (n=14,387)** |
| --- | --- | --- | --- |
| **Age at the most recent clinic visit; mean (SD)** | 44.0 (11.5) | 58.4 (9.7) | 48.6 (12.9) |
| **Sex (Female %)** | 7056 (71.8%) | 3211 (70.5%) | 10267(71.4%) |
| **Multiple sclerosis symptom Onset Age; mean (SD)** | 32.1 (10.0) | 33.0 (10.5) | 32.4 (10.2) |
| **Disease duration at the most recent clinic visit (Years); median [IQR]** | 10.0 [5.0 - 17.0] | 25.0 [17.0 - 33.0] | 14.0 [7.0 - 23.0] |
| **Most recent EDSS score; median [IQR]** | 1.5 [1.0 - 2.5] | 6.5 [4.5 - 7.5] | 2.5 [1.0 - 5.0] |

EDSS: Expanded disability scale status, IQR: Interquartile range, SD: Standard deviation.

### Decision tree classifier

All decision tree classifiers yielded similar accuracies ranging from 89.6% (95% confidence intervals (CI): 89.1 to 90.1) for the model containing the EDSS score, age, sex and disease duration to 89.3% (95% CI: 88.8% to 89.8%) for the model containing only the EDSS score and age. Given the simplicity of the latter and the similar accuracy between models, the model using most recently available EDSS score and age was chosen as the final decision tree classifier. The tree was pruned to its simplest state without any loss of accuracy, as shown in Table 2 and Figure 1.

**Table 2.**
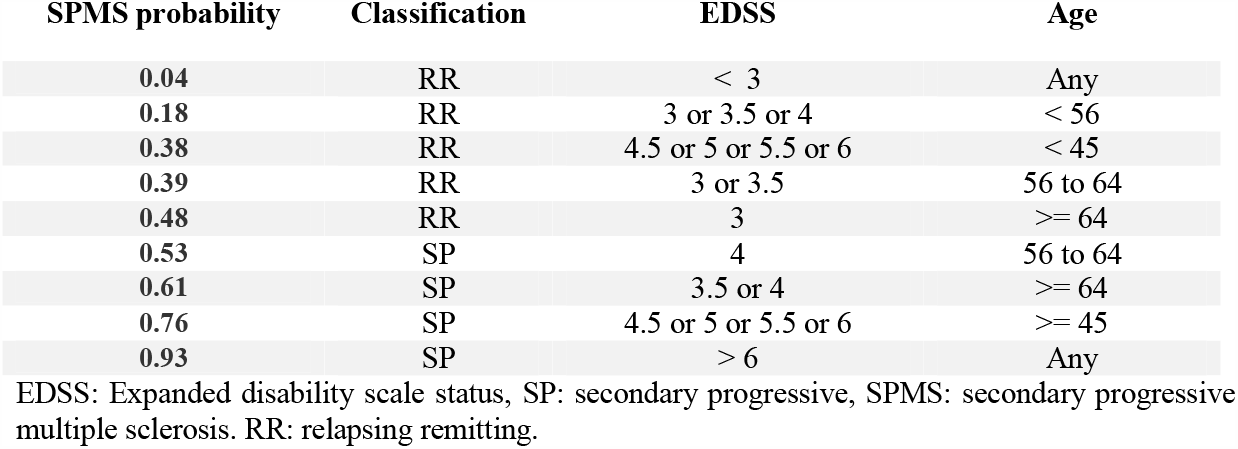
Full decision tree model and corresponding terminal node probabilities of SPMS. SPMS probability Classification EDSS Age

| SPMS probability | Classification | EDSS | Age |
| --- | --- | --- | --- |
| 0.04 | RR | < 3 | Any |
| 0.18 | RR | 3 or 3.5 or 4 | < 56 |
| 0.38 | RR | 4.5 or 5 or 5.5 or 6 | < 45 |
| 0.39 | RR | 3 or 3.5 | 56 to 64 |
| 0.48 | RR | 3 | >= 64 |
| 0.53 | SP | 4 | 56 to 64 |
| 0.61 | SP | 3.5 or 4 | >= 64 |
| 0.76 | SP | 4.5 or 5 or 5.5 or 6 | >= 45 |
| 0.93 | SP | > 6 | Any |

EDSS: Expanded disability scale status, SP: secondary progressive, SPMS: secondary progressive multiple sclerosis. RR: relapsing remitting.

**Figure 1:**
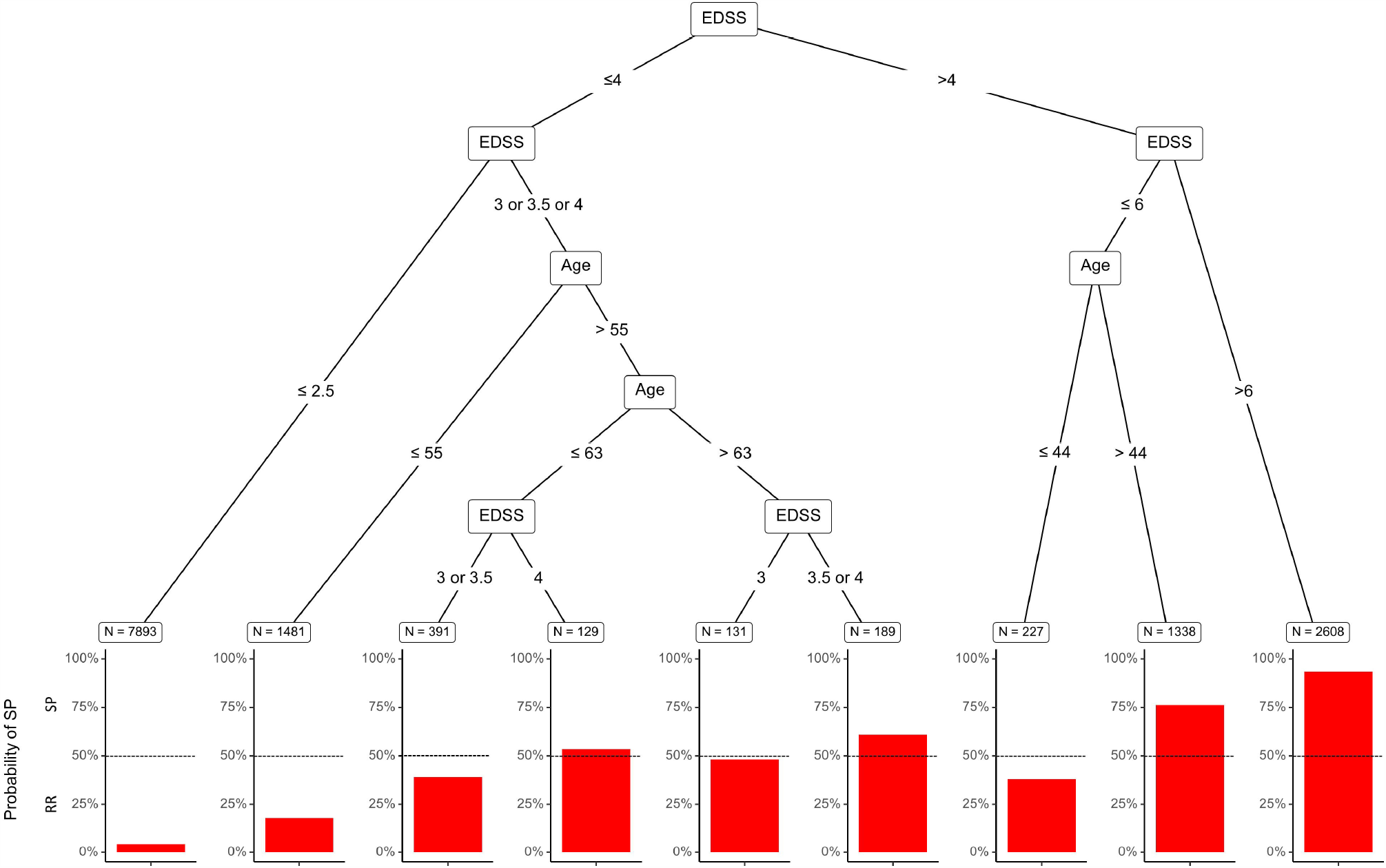
Pruned decision tree classifier based on a MS patient’s age and EDSS score. Terminal nodes are indicated by number of individuals and the bar length indicates probability of SPMS.

### Internal and external validation

The internal accuracy of the decision tree model, when constructed and tested on the Swedish cohort, was 89.3% (95% CI: 88.8% to 89.8%). The Canadian cohort included 5,431 relapsing-onset patients of whom 1,954 (36%) had transitioned to SP by end of follow-up. Mean age at the end of follow-up was 47 years (SD ±11.3) and median last available EDSS score was 3.5 (interquartile range: 4). When tested for validation accuracy in the Canadian cohort, the model was 82.0% (95%CI: 81.0% to 83.1%) accurate at determining the clinically assigned disease phenotype by an MS neurologist (Table 3).

**Table 3:** Accuracy of classifiers of SPMS, including sensitivity, specificity, Positive Predictive Value (PPV) and Negative Predictive Value (NPV)

| Classifier (and cohort) | N | Accuracy | Sensitivity | Specificity | PPV | NPV |
| --- | --- | --- | --- | --- | --- | --- |
| **Decision trees (Swedish cohort)** | 14,387 | 89.3% | 93.7% | 79.9% | 90.1% | 85.4% |
| **Decision trees (Canadian cohort validation)** | 5,431 | 82.0% | 89.8% | 71.4% | 81.2% | 83.5% |
| **MSBase algorithm (Swedish cohort)** | 14,387 | 77.8% | 76.6% | 85.5% | 97.2% | 35.9% |
| **Logistic Regression (Swedish cohort)** | 14,387 | 89.3% | 94.0% | 79.2% | 90.1% | 86.0% |
| **Neurologists (averaged)\*** | 100 | 83.7% | 92.8% | 53.2% | 88.0% | 66.7% |

\* Average accuracy of three neurologists examining full records of 100 patients. The Decision tree model classified 85 of these 100 correctly by comparison.

### Comparisons to MSBase algorithm

The MSBase algorithm achieved 77.8% (95%CI: 77.1% to 78.4%) classification accuracy when applied to the Swedish cohort. The MSBase algorithm is more conservative in assigning SPMS and achieved higher specificity and subsequently higher positive predictive value as compared with the decision tree classifier (Table 3). Characteristics of the patients misclassified by the decision tree and the MSBase algorithm as compared to the clinically assigned phenotype in the Swedish cohort are presented in Table 4. RR patients misclassified to SP in both approaches were generally older, with longer disease duration and high EDSS scores at the most recent clinic visit. SP patients misclassified to RR by decision tree had a significantly lower EDSS scores compared to clinically assigned SP patients. Misclassification of SP patients by the MSBase algorithm was mainly due to absence of confirmed progression.

**Table 4:** Characteristics of patients misclassified by the decision tree classifier and MSBase algorithm.

|  | Clinically assigned RR in the Swedish cohort |  |  | Clinically assigned SP in the Swedish cohort |  |  |
| --- | --- | --- | --- | --- | --- | --- |
|  | Clinically assigned RR (Reference phenotype) (n=9830) | Misclassified to SP |  | Clinically assigned SP (Reference phenotype) (n=4557) | Misclassified to RR |  |
|  |  | decision tree classifier (n=622) | MSBase algorithm (n=278) |  | decision tree classifier (n=915) | MSBase algorithm (n=2921) |
| **Age at the most recent clinic visit; mean (SD)** | 44.0 (11.5) | 55.7 (9.7) | 50.1 (11.3) | 58.4 (9.6) | 53.5 (10.4) | 58.0 (9.8) |
| **Sex (Female %)** | 7056 (71.8%) | 457 (73.5%) | 196 (70.5%) | 3211 (70.5%) | 660 (72.1%) | 2069 (70.8%) |
| **Multiple sclerosis symptom Onset Age; mean (SD)** | 32.1 (10.0) | 37.4 (11.5) | 33.8 (11.1) | 33.0 (10.5) | 33.8 (10.6) | 33.4 (10.6) |
| **Disease duration at the most recent clinic visit (Years); median [IQR]** | 10.0 [5.0 - 17.0] | 17.0 [10.0, 25.0] | 14.0 [9.0, 21.0] | 25.0 [17.0 - 33.0] | 18.0 [12.0, 25.5] | 23.0 [16.0, 32.0] |
| **Most recent EDSS score; median [IQR]** | 1.5 [1.0 - 2.5] | 5.5 [4.5, 6.5] | 5.0 [4.0, 6.0] | 6.5 [4.5 - 7.5] | 3.0 [2.0, 3.5] | 6.0 [3.5, 7.0] |

EDSS: Expanded disability scale status, IQR: Interquartile range, SD: Standard deviation. RR: relapsing remitting. SP: secondary progressive.

### Comparison to clinical evaluations by neurologists

Clinical evaluations by two neurologists on a randomly selected set of 100 patients when using only the most recent EDSS score and age were 79.0% and 87.0% accurate. The decision tree classifier was 85.0% accurate for these patients. Clinical evaluation of the same set of 100 patients but with complete clinical history by three neurologists had classification accuracies of 83.0%, 83.0% and 85.0% (average 83.7%), Table 3.

### Median time to SP conversion

From Kaplan-Meier curves, the median time to SP from birth, i.e., the age at which SP was reached was 60.1 (95%CI: 59.7 to 60.5) years for the decision tree classifier, 66.0 (95%CI: 65.4 to 66.8) years for the MSBase algorithm and 59.3 (95%CI: 58.8 to 59.7) years based on the clinical evaluations for the Swedish cohort (Figure 2). From Kaplan-Meier curves, the median time to SP from multiple sclerosis symptom onset was 26.3 (95%CI: 25.9 to 26.8) years for the decision tree classifier, 34.5 (95%CI: 33.6 to 35.6) years for the MSBase algorithm and 25.0 (95%CI: 24.5 to 25.5) years based on the clinical evaluation.

**Figure 1:**
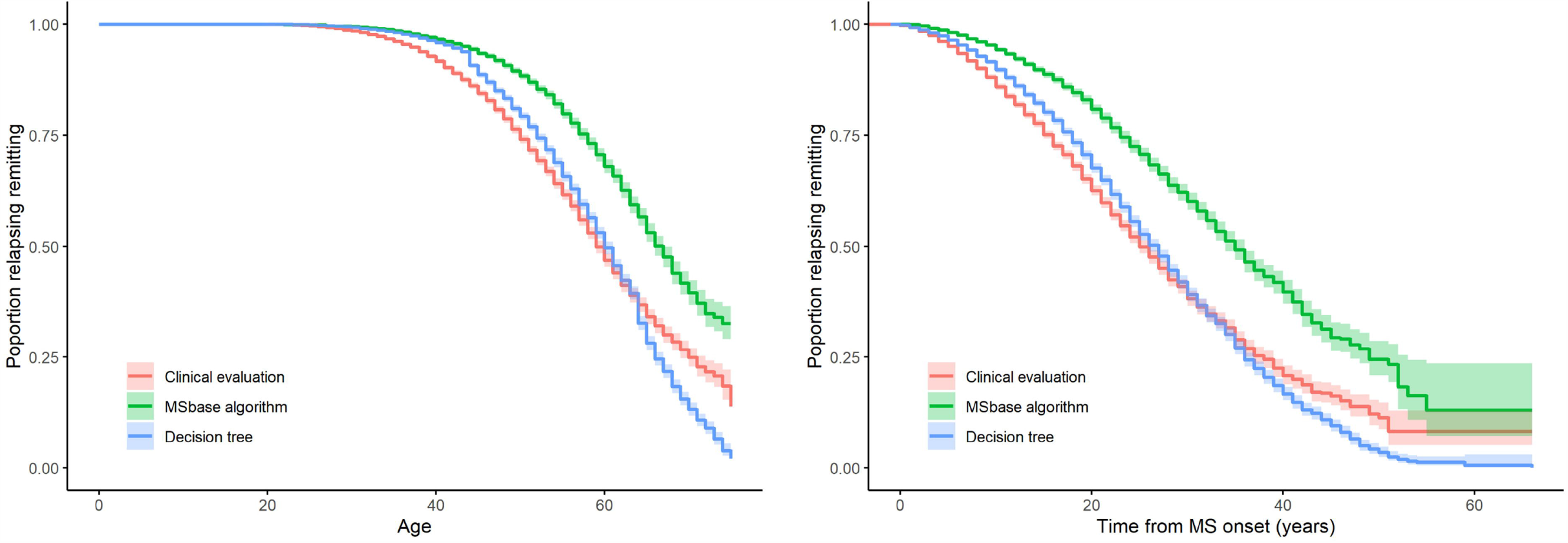
Kaplan-Meier estimate and 95% confidence intervals (coloured bands) of the various models based on age (left) and time from MS onset (right) in years at conversion to transition to SPMS (SP multiple sclerosis) (total RR-onset population n=13,712).

## Discussion

An accurate measurement of the probability of a patient having reached SP phase of multiple sclerosis could have great benefits for both clinical research and decision-making in clinical settings, especially given the hope that more disease-modifying drug options will be available to manage or delay SP phase of multiple sclerosis. The model presented uses a decision tree classifier to obtain highly accurate estimation of current clinical course, using only the patient’s most recent EDSS score and corresponding age.

Our decision tree classifier provides an objective assessment of multiple sclerosis phenotype (RR or SP). This may benefit multi-site studies, including multinational clinical trials because, at present, the determination of SP phase may vary between participating centers. Generally, applying the model to assign multiple sclerosis phenotypes may be less prone to personal or cultural biases when assigning SP status, however, certain limitations of the EDSS may still carry forward to this model. The model may also be of value to identify patients in need of more careful clinical evaluations or when clinical history is not available. Further, model based methods allow the identification of large and homogenous pools of data which can be used internationally, similar to the multiple sclerosis severity score (MSSS) and Age Related Multiple Sclerosis Severity (ARMSS) scores (Sawcer *et al*., 2005).

Authors of a 2016 study proposed an EDSS based objective measure of SP phase transition for an earlier and more consistent identification of SP patients (the MSBase algorithm) (Lorscheider *et al*., 2016). Although this proposed algorithm may increase the sensitivity and specificity of SP classification as compared to clinical evaluation and can also result in more consistent phenotype assignment, the method still relies on access to longitudinally collected data, including the FS sub-scores. Our current proposed decision tree classifier, which does not rely on longitudinal data, may therefore offer an alternative approach to phenotype determination in research studies since patients who are assumed to still be in the RR phase, but have not yet been reviewed clinically, can be accurately classified. As our classifier requires access to only limited amounts of clinical data, it may also have the benefit of increasing the pool of patients potentially eligible for analyses or inclusion in a study. However, reliance only on cross-sectional data may increase the misclassification rate, possibly more so for patients with a more stable disease. Consequently, the decision tree classifier yielded lower specificity than the MSBase algorithm which uses longitudinal data. The decision tree classifier incorrectly classified 622 of 9,830 RR (6.3%) patients as SP due to over reliance on cross-sectional data. These patients were on average five years older and had a significantly higher EDSS score (median: 5.5 vs. 1.5) at the time of their most recent assessment than the ‘general’ RR population who were determined to still be in the RR phase by the treating neurologist.

Ideally, our classifier, which is a form of supervised machine learning, could be compared against another objective measure of SPMS onset (e.g., a reliable bio- or imaging-marker). In the absence of such a marker, we compared to a neurologist-determined disease course which may be rather subjective. Clinical assignment of phenotype in our Swedish cohort is based on the collective contribution of hundreds of neurologists who typically follow their patients throughout their lives. Hence, a reasonably consistent and accurate classification of phenotypes by practicing neurologists for each of their patients is expected. This is despite that fact that each neurologist may classify their patients slightly differently with respect to EDSS and RR/SP status. A model trained on thousands of patients with different neurologists recording assessments may better generalize the differences than a single neurologist, resulting in increased consistency and accuracy. This may partly explain the lower than expected accuracy of classification by three neurologists in this work.

Although the model has high classification accuracy, caution must be exercised when interpreting an individual patient’s status in a clinical setting. For an individual patient, classifying their disease as having progressed to the SP stage may be unsettling as it can denote an irreversible decline in a patient’s underlying disease. Furthermore, this can trigger a discussion on DMT discontinuation, as many DMTs have limited effect on the disease course at SP phase. However, with potential for newly emerging DMT options in the treatment of SPMS (Hawker *et al*., 2009; Perrone *et al*., 2014; Kappos *et al*., 2018), this would likely mitigate DMT cessation and instead inform a potential treatment switch. Thus, this is a useful addition for providing information about the patient’s likely status during a clinical visit and to help with the neurologist’s decision-making in clinical settings regarding prognosis and treatment. Additionally, the decision tree classifier can serve as a marker to notify if the assigned RR course needs to be carefully revised.

Similar to the MSBase algorithm that showed lower accuracies in the Swedish cohort than the original cohort (Lorscheider *et al*., 2016), the accuracy of our decision tree classifier was expectedly slightly lower when applied to the Canadian cohort. This can be due to range of factors including; the different time periods between the Swedish & Canadian data (Canada being a more historical dataset), the differences in DMT availability during the different time periods and, differences in phenotype assignment during the different time points.

Nevertheless, the decision tree model constitutes an improvement based on not only improved accuracy, but also the extremely simple data requirements for which classification can be easily determined for patients during each clinical visit, as opposed to requiring clinical assessments over time for evidence of progression independent of relapse, and confirmatory EDSS scores. Simplicity of the decision tree facilitates its clinical and research utility.

## Acknowledgements

The authors wish to thank neurologists, nurses and multiple sclerosis patients in Sweden and Canada, as well as the Swedish Multiple Sclerosis Register for providing data for this study.

We gratefully acknowledge the BC MS Clinic neurologists who contributed to the study through patient examination and data collection (current members at the time of data extraction listed here by primary clinic): UBC MS Clinic: A. Traboulsee, MD, FRCPC (UBC Hospital MS Clinic Director and Head of the UBC MS Programs); A-L. Sayao, MD, FRCPC; V. Devonshire, MD, FRCPC; S. Hashimoto, MD, FRCPC (UBC and Victoria MS Clinics); J. Hooge, MD, FRCPC (UBC and Prince George MS Clinic); L. Kastrukoff, MD, FRCPC (UBC and Prince George MS Clinic); J. Oger, MD, FRCPC; Kelowna MS Clinic: D. Adams, MD, FRCPC; D. Craig, MD, FRCPC; S. Meckling, MD, FRCPC; Prince George MS Clinic: L. Daly, MD, FRCPC; Victoria MS Clinic: O. Hrebicek, MD, FRCPC; D. Parton, MD, FRCPC; K Atwell-Pope, MD, FRCPC. The views expressed in this paper do not necessarily reflect the views of each individual acknowledged.

## Funding

This study was funded by the Swedish Strategic Research Foundation, the Swedish Brain foundation and by the Swedish Research Council.

## Role of the funding source

The funders had no role in study design, data collection and analysis, decision to publish, or preparation of the manuscript.

## Competing interests

RR has nothing to disclose.

AM is supported by the Margaretha af Ugglas Foundation

KF received an unrestricted research grant from Biogen, NeuroFonden and Neuroföbundet. KF has received travel compensation for lectures for Novartis, Biogen, TEVA, Almirall and Merck.

VK received financial support from Stockholm County Council and Biogen’s Multiple Sclerosis Registries Research Fellowship Program. VK has also received an unrestricted grant from Biogen and a project grant from Novartis.

JH received honoraria for serving on advisory boards for Biogen and Genzyme and speaker’s fees from Biogen, Novartis, Teva and Sanofi-Genzyme. He has served as P.I. for projects sponsored by, or received unrestricted research support from, Biogen, Sanofi-Genzyme and Novartis. His multiple sclerosis research is funded by the Swedish Research Council and the Swedish Brain Foundation.

HT is the Canada Research Chair for Neuroepidemiology and Multiple Sclerosis. Current research support received from the National Multiple Sclerosis Society, the Canadian Institutes of Health Research, the Multiple Sclerosis Society of Canada and the Multiple Sclerosis Scientific Research Foundation. In addition, in the last five years, has received research support from the UK MS Trust; travel expenses to present at CME conferences from the Consortium of MS Centres (2018), the National MS Society (2016, 2018), ECTRIMS/ ACTRIMS (2015, 2016, 2017, 2018, 2019, 2020), American Academy of Neurology (2015, 2016, 2019). Speaker honoraria are either declined or donated to an MS charity or to an unrestricted grant for use by HT’s research group.

EK is supported through research grants from the Canadian Institutes of Health Research and the Multiple Sclerosis Society of Canada. In addition, during the last five years, she has received travel expenses to give presentations, or attend CME conferences, from ACTRIMS, ECTRIMS, and the MS Society of Canada.

PB and FZ have nothing to disclose.

JL received research support from Innosuisse – Swiss Innovation Agency, research grants from Biogen and Novartis and honoraria for serving on advisory boards from Roche and Teva.

## AUTHOR CONTRIBUTIONS

RR designed the study, analyzed and interpreted the data, wrote and revised the paper for important intellectual content.

KF collected the data and revised the manuscript for important intellectual content. VD collected the data and revised the paper for important intellectual content.

JH interpreted the data and revised the manuscript for important intellectual content.

AM designed the study, analyzed and interpreted the data, wrote and revised the paper for important intellectual content.

EK interpreted the data and revised the manuscript for important intellectual content. HT interpreted the data and revised the manuscript for important intellectual content.

FZ helped with the analysis, interpreted the data, and revised the paper for important intellectual content.

JL helped with the analysis, interpreted the data, and revised the paper for important intellectual content.

PB helped with the analysis, interpreted the data, and revised the paper for important intellectual content.

## Notes

### Author Declarations

Ethical permission for the study was granted by the Stockholm Regional Ethical Committee and the University of British Columbia's Clinical Research Ethics Board.

